# Improving Risk Assessment of Miscarriage during Pregnancy with Knowledge Graph Embeddings

**DOI:** 10.1101/2020.06.04.20122150

**Authors:** Hegler C. Tissot, Lucas A. Pedebos

**Affiliations:** University College London; Secretaria Municipal de Saude de Florianopolis

**Keywords:** Machine learning, Knowledge based systems, Risk analysis, Health information management, Public healthcare

## Abstract

Miscarriages are the most common type of pregnancy loss, mostly occurring in the first 12 weeks of pregnancy due to known factors of different natures. Pregnancy risk assessment aims to quantify evidence in order to reduce such maternal morbidities during pregnancy, and personalized decision support systems are the cornerstone of high-quality, patient-centered care in order to improve diagnosis, treatment selection, and risk assessment. However, the increasing number of patient-level observations and data sparsity requires more effective forms of representing clinical knowledge in order to encode known information that enables performing inference and reasoning. Whereas knowledge embedding representation has been widely explored in the open domain data, there are few efforts for its application in the clinical domain. In this study, we discuss differences among multiple embedding strategies, and we demonstrate how these methods can assist on clinical risk assessment of miscarriage both before and specially in the earlier pregnancy stages. Our experiments show that simple knowledge embedding approaches that utilize domain-specific metadata perform better than complex embedding strategies, although both are able to improve results comparatively to a population probabilistic baseline in both AUPRC, F1-score, a proposed normalized version of these evaluation metrics that better reflects accuracy for unbalanced datasets.

## I. Introduction

OVER 200 million women become pregnant each year worldwide, and more than one fourth of these are estimated to end in pregnancy loss, even in developed regions as shown by some estimates [1], [2]. A high-risk pregnancy threatens the health (or life) of both mother and fetus, requiring specialized care. Some pregnancies become high risk as they progress, whereas some women are at increased risk for complications even before they get pregnant for a diverse set of reasons, including (a) existing health conditions (e.g. high blood pressure, diabetes, or being HIV-positive), (b) overweight and obesity, that can lead to other complications, such as high blood pressure, preeclampsia, and gestational diabetes, and (c) multiple births, and (d) young or old maternal age, in the latter mostly due chromosomal abnormalities [3],[4].

Pregnancy risk assessment aims to quantify evidence on risks and risk-outcome associations in order to reduce infant and maternal morbidity by influencing maternal behaviors before, during, and immediately after pregnancy. Maternal morbidities includes any condition that is attributed to or aggravated by pregnancy and childbirth which has a negative impact on the woman’s wellbeing and/or functioning, such as hemorrhage, eclampsia, sepsis, complications of obstructed labor, and miscarriage [5].

Miscarriages are the most common type of pregnancy loss and being both physically and emotionally painful. Mostly occurring in the first 12 weeks of pregnancy, they go usually unnoticed most of the time due to the delay in the perception of pregnancy by most women. However, there are known factors of different natures related to the increase in the occurrence of spontaneous abortions, such as abnormal fetus development, use of drugs that interfere with pregnancy, diabetes mellitus, and previous kidney problems [6].

Although the number of high quality health studies has made it possible to develop several evidence-based guidelines throughout the last years, there is still a lack of refinement in the use of case-oriented data in the everyday clinical practice. Personalized medicine has the potential of switching health-care standards from common guidelines to solid computational models based on data obtained for an individual patient in order to improve diagnosis, treatment selection, and health system efficiency. This is a new frontier health professionals and decision makers are facing, in which computational methods based on large sets of patient-centric data could become a new standard for clinical evaluation and prediction [7], [8].

Due to the large number patient-level observations and data sparsity, a new machine learning paradigm based on Knowledge Graphs (KG) recently emerged of considering flexible rather than fixed sets of features used to describe each individual [9]. The flexible schema used to store information in the form of relationships (edges) between entities (nodes) enables the interactions of multiple clinical factors to be analyzed more effectively, moving from population-based approaches to patient-centric predictions [7], [8].

Many methods for learning knowledge embedding representation (KER) have been developed. KER operates by(i) learning a low-vector representation of KG constituents (nodes and edges) that preserves the graph inherent structure and the semantics of different types of associations between entities, and (ii) exploiting low-rank latent structure in the data to encode known information that enables inference. Hence, KER models fit for performing analytical and predictive tasks that verify and infer multiple types of interactions between entities, as when representing complex patient-centric clinical knowledge [10].

In previous KER studies focused on open-domain tasks using more sparse data, although benchmark evaluation has steadily improved, approaches are complex and the latent structure still remains unexplained [11]. Recent studies have shown that more simple approaches that incorporate ontological constraints favoring embedding quality are able to speed up the training process and achieve better performance when evaluated in similar benchmark tasks for more dense domain-specific clinical datasets [12], [13]. We aimed to test whether KER approaches could assist on clinical risk assessment. We test the ability of embedding approaches on both capturing the semantic correlations from multi-relational categorized data and performing inference, and we demonstrate that even simple embedding approaches that utilize domain-specific metadata are able to improve the risk evaluation both before and during the earlier pregnancy stages.

## II. Materials & Methods

### A. Motivation

Although global risk assessment can provide an overall picture for a given comorbidity, some populations can be under-represented and geo-related factors disregarded. Thus, the use of regional rather than national data is justified. Locally led research can support the evidence to understand the temporal changes in risk-attributable components (e.g. population growth, changes in population age, and changes in exposure to behavioral metabolic) when primary health care systems provide patient-centric individualized historical series coupled with a controlled and high-quality stable scenario that supports performing data analysis and designing more representative comparable risk assessment models risks [14].

In 2018, the Brazilian Ministry of Health elected the municipality of Florianópolis as the best primary health care capital within the National Program for Improving Access and Quality in Primary Care (PMAQ) [15], and also ranked it among the top-3 cities with the best Performance Index of the Unified Health System (ID-SUS). Moreover, Florianópolis has been using electronic medical records over the last 20 years, from which over 80% of the health network data being stored in digital format since 2008. *InfoSaude* [16],[17] is an Electronic Health Record (EHR) system created to manage and track medical records used to meet the needs of Florianópolis’ 75 public health centers, integrating patient EHRs with multiple information structures, such as distinct types of care, pregnancies, procedures performed on each patient, applied vaccines and drug prescriptions.

Analysis of miscarriage reports from *InfoSaude* (Fig. 1) shows that the distribution of miscarriages regarding maternal age has been changing over the last decade. The proportion of cases in women under 25 decreased from 40% to 27% between 2008 and 2018, whereas the same proportion increased from 12% to 38% in the same period for women over 35. The former reflects some of the public health policies associated with specific early-aged pregnancy programs implemented over the last 15 years. The latter, however, evinces the odds in healthcare efforts and priorities. Finally, pregnancy losses in [25–35]-year-old women tend to remain proportionally constant, with a slight increase in the period that coincides with spread of Zika virus in Brazil (2015-2016), with an estimated birth reduction of 7.78% response to Zika virus-associated microcephaly rate [18].

**Fig. 1.**
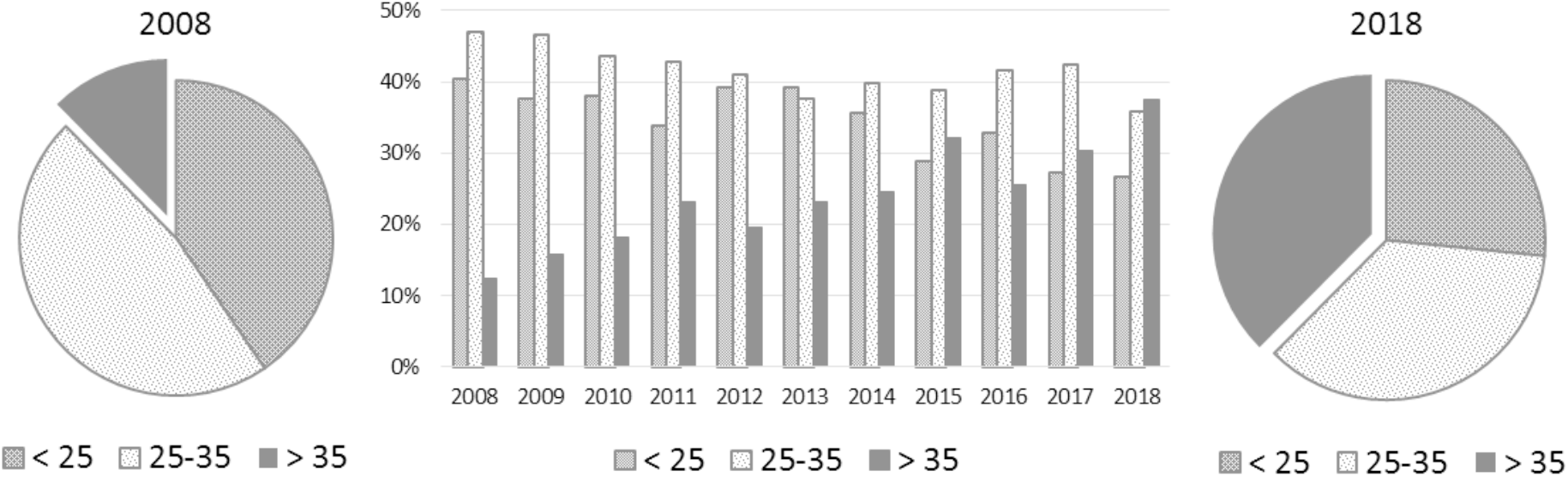
2008-2018 distribution of miscarriages per maternal age: (a) in 2008, miscarriages in women over 35-year-old represented only 12% of the cases; (b) over the years, the proportion of miscarriages in older women consistently increases; (c) in 2018, miscarriages in women over 35-year-old represented almost 40% of the cases.

Changes in the patterns of socioeconomic factors can also play important role on significantly increasing the number of pregnancies in older women [19]. Moreover, pregnancy after age 35 that makes certain complications more likely to directly reflect an expecting increasing number of high risk pregnancies. However, within the *InfoSaude* cohort, there are several factors contributing to reduce the proportion of high-risk pregnancies in the last decade, which include (a) improvements in the quality of primary care services and healthcare programs, (b) use of well-defined health protocols leading to better control of chronic illnesses, and (c) adoption of smoking reduction programs. The latter, for example, allowed a reduction in the proportion of female smokers from 15.8% to 8.6% in the period between 2006-2016, as reported in a study that estimated sociodemographic frequency and distribution of risk and protective factors for chronic diseases in the capitals of the 26 Brazilian states and the Federal District in 2016 [20]. Indeed, Fig. 2 shows that the percentage of pregnancies flagged as ‘high risk’ has decreased from 6.5% to less than 3.0% within a 10-year period. However, the corresponding number of cases of miscarriages has increased from 3.7% up to 6.2% in the same period, which raises an alert on how pregnancy risk assessment protocols have been effectively applied.

**Fig. 2.**
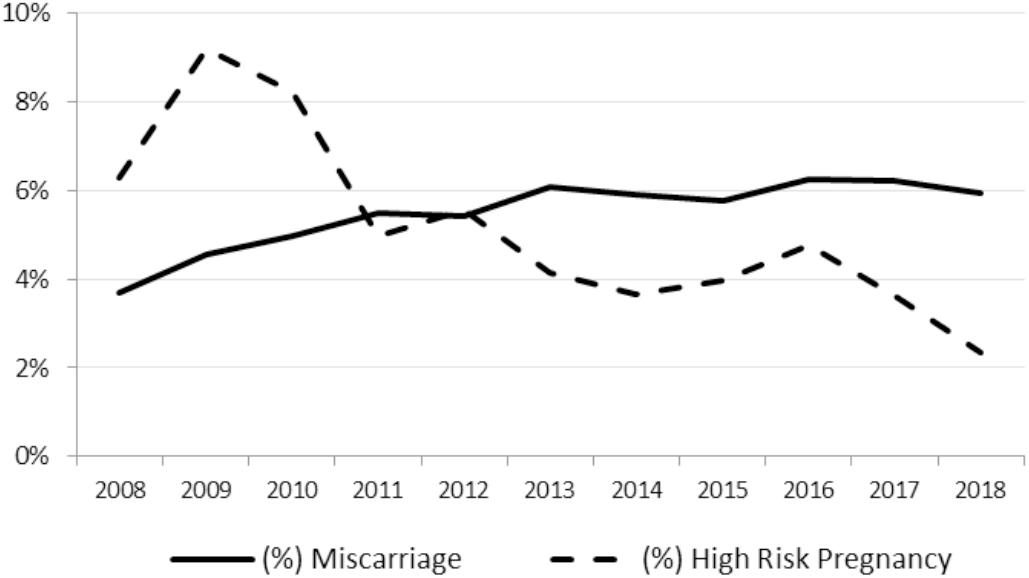
Proportion of miscarriages vs. high-risk pregnancies contrasted over the years (2008-2018)

### B. Dataset

In this work, we combined structured and unstructured data extracted from the *InfoSaude* system regarding 24,877 pregnancies occurring in the period from 2010 to 2015 in order to evaluate how KER approaches could better assist on performing risk assessment of miscarriage during pregnancy.^1^

Structure data covering most of the demographic and clinical history before and during pregnancy was coupled with additional unstructured information relevant to this use-case, the latter extracted from short notes about history of allergies, infections and other clinical conditions. Such notes are written by physicians usually limited to 50-100 characters. Very simple regular expressions were able to extract the most common facts as well as the language signals for eventual negations. Finally, a hybrid phonetic similarity algorithm [17], [21] was used to find and merge misspellings, positively identified and manually checked for about 1.5% of the mentions in each considered sets of substances and infections.

Infections during pregnancy are a frequent cause of morbidity and mortality among mothers, fetuses and neonates, leading to miscarriages, preterm births or fetal deaths, and fetal malformations and growth restrictions. Infections can be asymptomatic or manifest symptoms similar to those in non-pregnant individuals, and their risk of causing pregnancy outcomes depends on factors such as (a) prevalence of infections in the population, (b) socioeconomic and cultural factors, and (c) health and healthcare-seeking behavior [22]. However, we found there is not a systematic way of collecting information for several features taken into account in this study, e.g. regarding history of infections within the analyzed population,in which only 2.5% of the pregnancies have some history of infection associated in their corresponding EHR.

For each pregnancy, we collected a set of features subsequently mapped as relations in the KG (coverage % below is given to each feature in which data is not available for all cases):

- <u>Demographic data</u>: year when pregnancy started, age in years on the corresponding first day of last menstrual period (LMP), marital status, ethnicity, education level, neighborhood, profession (36.7%) and occupation (7.1%), country of birth (1.8% not from Brazil), and assigned local health office.
- <u>Clinical history</u>: weight (39.5%), height (34.3%) and BMI (25.6%) on LMP, the most common known infections (2.5%) ∈ {HIV, Bronchitis, Candidiasis, Hepatitis, Pyelonephritis, Syphilis, Toxoplasmosis, Urinary, Vaginitis, Vaginosis}, the most common known drug allergies (5.1%) ∈ {AAS, Iodine, Acetaminophen, Amoxicillin, Ampicillin, Benzetacil, Buscopan, Diclofenac, Dipyrone, Metoclopramide, Nimesulide, Penicillin, Plasil, Sulfa}, and other known clinical conditions (12.2%) ∈ {alcohol abuse, smoker, anemia, hypertension}.
- <u>Prescriptions</u>: each medication is identified by the generic name of the substance used – a total of 170 substances are being considered in this dataset.
- <u>Diagnoses</u>: each diagnosis is given by the corresponding ICD-10 code coupled with the corresponding appointment’s service group (an administrative concept used within the *InfoSaude* system) and the physician’s medical specialty.
- <u>Procedures</u>: each procedure is coupled with the corresponding nurse’s or physician’s medical specialty.

The time window we collected data regarding prescriptions, diagnoses, and procedures is given by: (a) a one-year history before the LMP split into each one of the preceding quarters (Q1-Q4), and (b) the first 24-week pregnancy period split into 3 periods of 8 weeks each. Each split adds variations in the names of each relation in order to somehow incorporate a temporal context within the dataset, so that the same drug prescribed right before pregnancy and almost one year before pregnancy will be given by distinct relations, `Prescription_Q1` and `Prescription_Q4` respectively. The number of triples in each pre- and post-LMP period is given in Table I, and Fig. 3 shows an example on how a pregnancy is represented as a set of triples (subject, relation, object) in the resulting dataset.

**Table I:** Statistics of training and test sets.

| Stats | Training Set | Test set |
| --- | --- | --- |
| Years | 2010-2014 | 2015 |
| Pregnancies | 20,201 | 4,676 |
| Triples (pre LMP) |  |  |
| Demographics | 166,436 | 37,266 |
| Clinical history | 63,745 | 14,376 |
| Prescriptions | 103,010 | 20,882 |
| Diagnoses | 143,667 | 31,669 |
| Procedures | 54,520 | 13,385 |
| Triples (post LMP) |  |  |
| Weeks 01-08 | 107,870 | 25,380 |
| Weeks 09-16 | 197,831 | 45,403 |
| Weeks 17-24 | 191,133 | 45,226 |

**Fig. 3.**
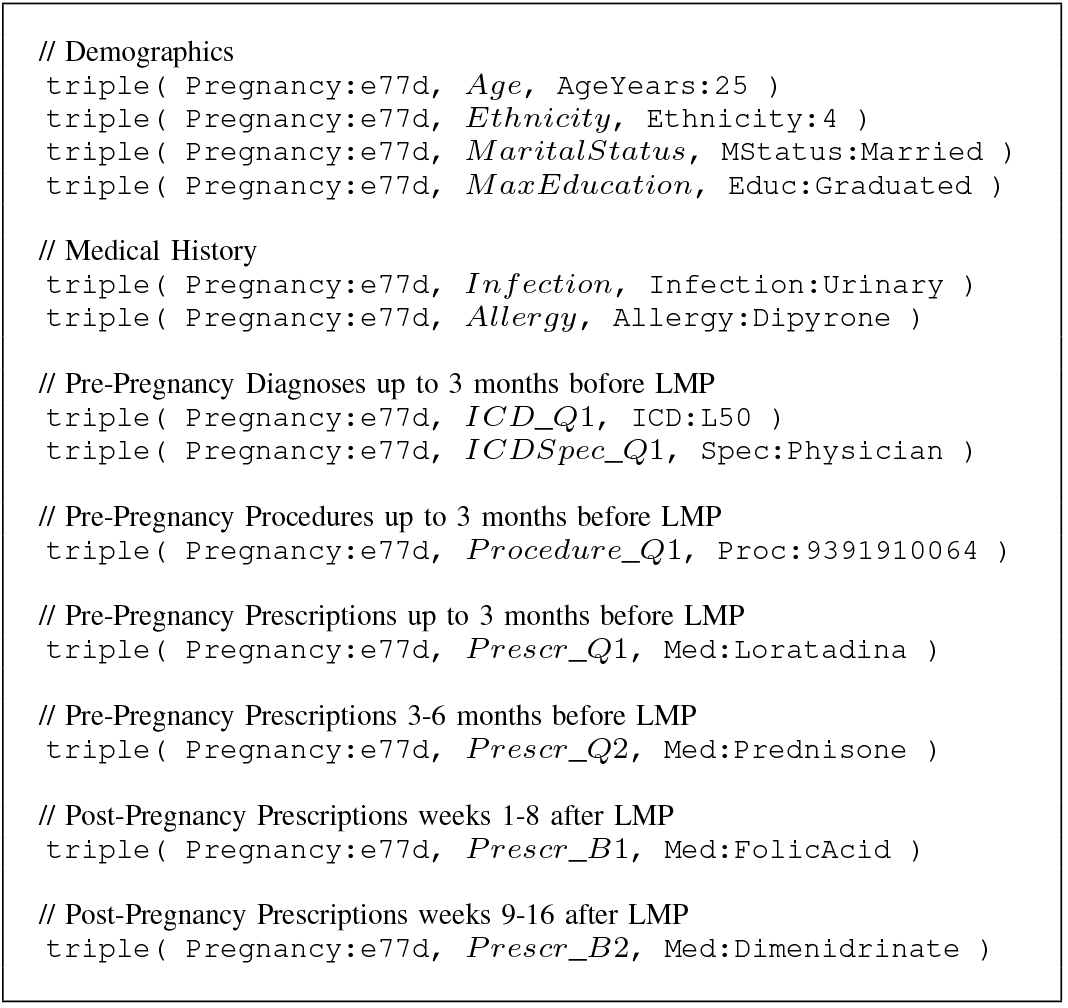
Example of pregnancy represented by a set of triples extracted from the KG dataset – patient ID was synthetically generated, and triples in this example were randomly selected from multiple patients in order to avoid identification.

Training and test sets were evenly and cumulatively split into: (a) ‘LMP’ includes demographic data, clinical history and the one-year history of prescriptions, diagnoses and procedures preceding the first day of the LMP for each confirmed pregnancy; (b) ‘Weeks 01-08’ adds prescriptions, diagnoses and procedures extracted from the first 8 weeks following LMP; (c) ‘Weeks 09-16’ adds prescriptions, diagnoses and procedures extracted from weeks 9-16 following LMP; and (d) ‘Weeks 17-24’ adds prescriptions, diagnoses and procedures extracted from weeks 17-24 following LMP. This setup allowed us to assess risk in four different points in time, starting from the beginning of pregnancy, and going through the first 5-6 gestational months.

Although the miscarriage flag labels are not part of the datasets, we found data signals in the KG (e.g. ICD-10 diagnosis codes corresponding to miscarriage) that could potentially influence scores and bias the results favoring positive labeled cases in later pregnancy stages. Thus, pregnancies in which the miscarriage occurred in any of the three periods of 8 weeks after LMP were removed from all subsequent test sets since they happened.

### C. Knowledge Embedding Representation

Knowledge Graphs (KGs) are a widely used representation for multi-relational data, comprising entities (nodes) and relations (edges) that provide a flexible structured schema adapted for both open- and domain-specific knowledge bases. Embedding representation methods are able to (a) learn and operate on the latent feature representation of the KG constituents and on their semantic relatedness, (b) efficiently handle data sparsity and inconsistency, such as missing or multiple inconsistent values for ‘one-to-one’ relations, and (c) enhance subsequent machine learning applications.

TransE [23] is a well-known translational approach that uses simple assumptions to achieve considerably accurate and scalable results, proved to be effective and efficient embedding model [24]. Several enhanced models (e.g. TransH [25], TransR [26], Rescal [27], HolE [28], and Complex [29]) were proposed to address TransE’s supposed flaws on representing ‘many-to-many’ relations. These methods attempt to couple TransE with more complex relation-specific representation of different data cardinality (e.g. projection matrices) in order to improve performance on the link prediction benchmark task (also known as knowledge graph completion). However, we show in previous work [13], [30] that these models do not necessarily provide a good embedding representation, in which similar entities are expected to be found in small clusters along the resulting embedding space. Indeed, embedding methods in general tend to report weak performance [11], [31].

Besides, we observed other well-known state-of-the-art models in open-domain benchmark tasks, such as Rescal, HolE and Complex, can take up to 30-50 times more CPU resource to perform training, whereas is still unclear the ability of these models to properly embed domain-specific knowledge representation. Experimental results testing these models with clinical datasets showed that, even after extending the training process over the usual 1000 learning epochs, the standard embedding evaluation protocol still shows very poor accuracy for dense clinical KGs. Moreover, traditional embedding systems are designed to tackle the evaluation task instead of favoring embedding quality, and the resulting embedding representation tends to cluster entities by their use as head or tail in each relation instead of their semantic relatedness. As a side-effect, groups of entities (similar type) are formed in such a way that makes the link prediction task hard, even when types are not explicitly given as part of the KG metadata.

Given the known flaws of translational models and their limitations on using more dense domain-specific datasets, HEXTRATO [12] emerged a translational embedding approach that couples TransE with a set of ontology-based constraints to learn representations for multi-relational categorized data, originally designed to embed biomedical- and clinical-related datasets. HEXTRATO improves the translational embedding from TransE and other enhanced models, achieving great performance, even in very low *k*-dimensional spaces (*k* < 100), without necessarily adding complex representation structures within the model training process, such as those used in TransE-based enhanced models.

Link prediction (LP) is a traditional evaluation protocol used during training when learning knowledge embedding representation. However, in previous work [13], we evaluated whether LP accurately reflects the quality of the resulting embeddings for multi-relational data, showing that more complex embedding representation approaches used to improve KG completion do not reflect into embedding quality. Thus, we extended HEXTRATO with additional features in order to mimic enhanced TransE models that use projection matrices, but still keeping the idea of using set of independent type-associated hyperspaces in order to project each entity belonging to the same type. We developed an embedding framework that allows implementing and contrasting multiple embedding representation methods,^2^ and we used this framework to mimic and simulate what most of the methods enhancing TransE use as embedding strategy, coupling the vectors resulting representation of each relation from TransE with projection matrices that aim to improve accuracy in traditional benchmark datasets. The four approaches taken into account along our experiments are described as follows:

- TransE: the original TransE approach using a vector representation for each relation;
- TransE+: a projection matrix is added to the embedding representation of each relation, mimicking some of the TransE enhanced approaches (e.g. TransH and TransR);
- KRAL: this is the original HEXTRATO approach, in which independent hyperspaces are used for each type of entity, and similarly to TransE, it uses a vector representation for each relation;
- KRAL+: similarly to TransE (Ext), a projection matrix is added to the embedding representation of each relation.

### D. Risk Assessment

In order to generate a risk score for each pregnancy we look at the embedding neighborhood of each pregnancy. Within the resulting 64-dimensional embedding space with radius 1.0, we use multiple maximum *L*_2_-norm radiuses ranging from 0.125 to 0.5 to calculate the percentage of neighbors labeled as positive to miscarriage (32 distinct values in total). Then, we compare the resulting F1-score from each neighborhood radius.

Very small radiuses can eventually find no neighbor entities, whereas very high radiuses will consider too many entities, so that the resulting ratio of positive cases would approximate the overall percentage of miscarriages in the entire dataset. Within our experiments, we found the risk assessment reached best F1 scores when using a radius of 0.321025.

We used two baselines for miscarriage risk assessment. Firstly, a ‘Random’ risk score representation is created in order to serve as primary baseline. Then, a population-based approach uses pregnancy features to provide a probabilistic score, presented as ‘Prob(F)’ within our results.

Prob(F) takes into account all *P* (*f, v*) supported by a minimum number of cases. *P* (*f, v*) corresponds to the probability of the target label (a miscarriage in this case) happening when a feature *f ∈ F* has a value *v ∈ V* (*F*). For example, *P* (*AgeYears*, 45+) = 0.2302 is the probability of a miscarriage happening in a patient over 45 years-old, whereas *P* (*PrescriptionPrevQ*1*, Fluoxetine*) = 0.1135 is the probability of a miscarriage happening for a patient prescribed with Fluoxetine in the quarter preceding LMP. We only consider features supported by a minimum number of 86 cases, which is the minimum sample size needed to reach a confidence level of 95% when resulting scores are within *±*5% of the measured/surveyed value, for a training population of 20,201 cases and the proportion of cases labeled as miscarriages is 5.91%. For each pregnancy, the probabilities all known pairs (*f, v*) fitting the previous condition are average in order to provide a final risk assessment score.

### E. Evaluation

High class imbalanced data and skewed data distributions are naturally observed in many real-world scenarios, such as in medical diagnosis-related tasks, in which the majority of the cases represent healthy patients as the negative class. Imbalanced data poses further challenges related to bias towards the majority class, even ignoring the minority target class in extreme cases, typically (a) under-classifying the minority group, and (b) misleading result with high scores that incorrectly indicate good performance [32].

Receiver operating characteristics (ROC) curve is a usual assessment to contrast true vs. false positive rates. However, ROC curves can exhibit unduly optimistic results, as the false positive rate (FPR) is less sensitive to changes in false positive (FP) as longer the size of the negative class is predominant [33]. Precision-Recall (AUPRC) curves are recommended be used for imbalanced data instead [33].

In our dataset, the ratio between negative and positive cases is ≈ 16.91 (5.91% of positive cases), which means there are almost 17 negative cases for each positive one, which increases the probability of more false positive cases happening along the evaluation, worsening the results due to the unbalanced distribution of the target label. Thus, in addition to presenting the original AUPRC and F1 scores, we also present weight-adjusted scores relying on a normalized version of false positive (FP) cases, which better approximates the scores to a balanced-similar scenario, potentially facilitating further analysis and comparison of embedding approaches on multiple datasets. F_*norm*_-score uses FP_*norm*_ instead of FP:

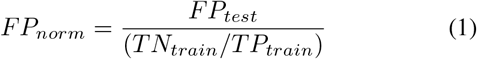

where FP is the number of false positive cases in the test set,

TN_*train*_ is the number of true negative cases in the training set, and TP_*train*_ is the number of true positive cases in the training set. FP_*norm*_ is used as a replacement to the original FP when calculating AUPRC_*norm*_ and F_*norm*_ scores.

## III. Results

The *InfoSaude* system reports 270 cases of miscarriage out of 4,676 pregnancies in which LMP occurred in 2015 (5.77%), corresponding to the test set in our experiments. Our method attempts at embedding a pregnancy knowledge graph in order to find clusters of similar patients that can support the decision of establishing a high risk pregnancy.

In table II we present AUPRC, F1 and F_*norm*_ scores for each approach evaluated and Fig. 4 compares all approaches regarding AUPRC in each pregnancy phase, starting at the beginning of pregnancy (LMP), followed by each period of 8 weeks after LMP.

**Fig. 4.**
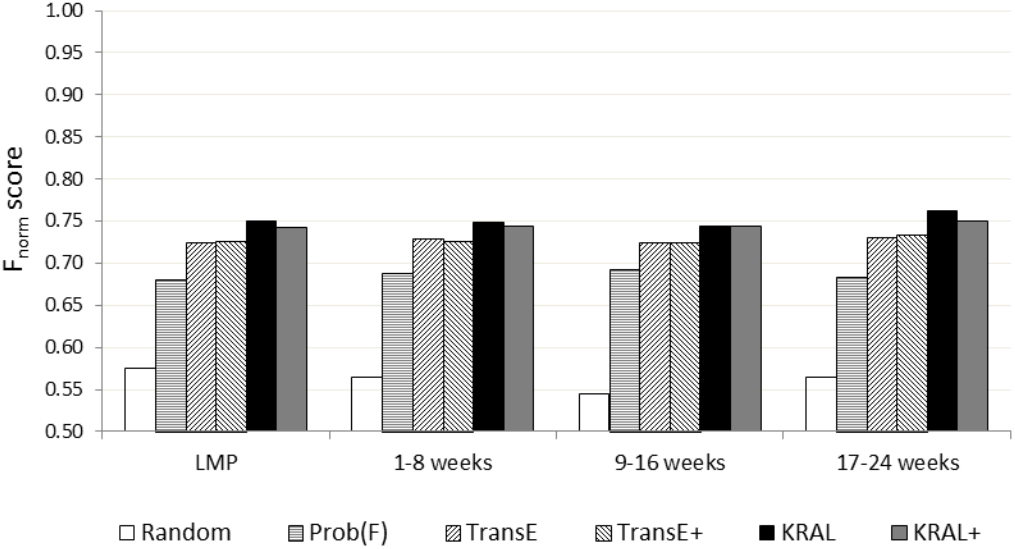
Overall results: Weighted F1_*norm*_ scores for translational embedding and baseline models.

**TABLE II.**
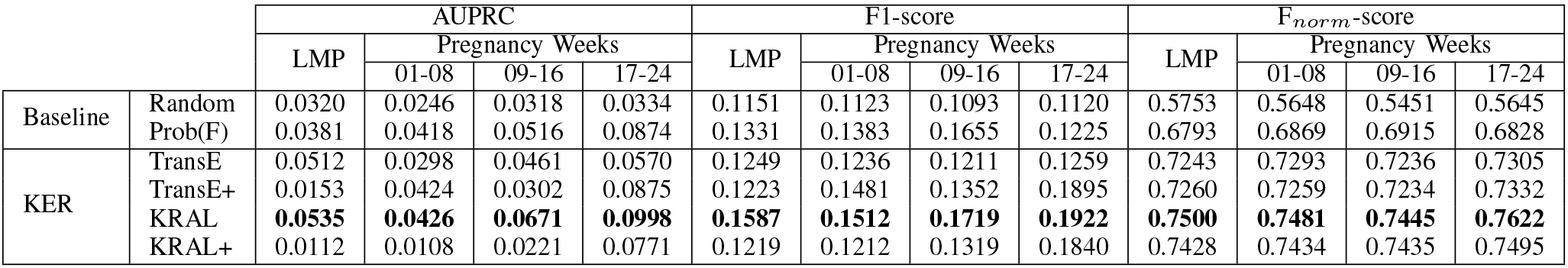
AUPRC, F1 AND F_*norm*_ SCORES.

The random approach performs as expected in a binary classification task, with F_*norm*_ scores close to 0.5 along all pregnancy phases. Then, Prob(F) presents as an efficient candidate as a baseline method for risk assessment–as long as combining the probabilities of miscarriages (target labels) occurring in each pair (*f, v*) of features *f* and their corresponding values *v*, it performs F_*norm*_ scores in the range of ≈ [0.67,0.69]. Finally, translational embedding approaches are able to preserve the graph inherent structure and the semantics of different types of associations between entities, and even though F_*norm*_ scores improvement is relatively low regarding Prob(F), embedding methods are able to support analytical and predictive tasks that require inferring multiple types of interactions between entities that represent complex patient-centric clinical data. KRAL performs slightly better than other embedding strategies, reaching F_*norm*_ scores ≈ 0.75.

In addition to analyzing model performance along the evaluation task, we also compare differences between each resulting model regarding statistical significance. We used a dependent sample paired Sample T-Test to determine whether the mean difference between two sets of observations can be considered statically significant. Table III presents the resulting *p*-value and *t* when comparing two models, considering significance level (alpha) = 0.05, H_0_ (A = B) is rejected when p-value < alpha, and |*t*| accepted range [−1.9605: 1.9605].

**Table III.** Significance tests between resulting risk assessment models.

| Approaches |  | Split | p-value | t |
| --- | --- | --- | --- | --- |
| Random | Prob(F) | LMP | -3.5527e-15 | 11.9903 |
|  |  | W(01-08) | 0.0000e-00 | 33.3973 |
|  |  | W(09-16) | 1.8874e-15 | -15.1822 |
|  |  | W(17-24) | 0.0076e-00 | -2.6700 |
| Prob(F) | TransE | LMP | 3.1978e-10 | -6.3025 |
|  |  | W(01-08) | 0.0000e-00 | -138.7982 |
|  |  | W(09-16) | 0.0000e-00 | -110.0519 |
|  |  | W(17-24) | 0.0000e-00 | -97.3523 |
| TransE | TransE+ | LMP | 0.2691e+01 | 1.1052 |
|  |  | W(01-08) | 0.3109e+01 | 1.0853 |
|  |  | W(09-16) | 0.8873e+01 | 0.1418 |
|  |  | W(17-24) | 0.1998e+01 | -1.1492 |
| KRAL | KRAL+ | LMP | 4.44089e-16 | 77.8029 |
|  |  | W(01-08) | 1.11022e-16 | -46.2310 |
|  |  | W(09-16) | 1.11022e-15 | 12.3938 |
|  |  | W(17-24) | 5.55112e-16 | -34.9729 |
| KRAL | TransE | LMP | -2.22045e-16 | -27.0375 |
|  |  | W(01-08) | -1.05386e-15 | -19.2891 |
|  |  | W(09-16) | -1.04290e-15 | -16.2562 |
|  |  | W(17-24) | -2.22045e-16 | -19.0259 |

F1_*norm*_ scores clearly show the improvement in performing risk analysis when moving from Random to Prob(F), and from the latter to embedding methods. However, it is still unclear whether more complex translational methods are able to improve the representativeness of a resulting embedding representation, as long as previous works have been mostly focused on link prediction as an evaluation task instead of assessing the quality of the resulting embedding models [13].

In open-domain data, the addition of projection matrices to TransE (generally represented in our experiments by TransE+) has not been able to add any significant signal that could be used to improve the evaluation task, and there was no significant statistical differences observed when comparing these models to the original TransE. Similarly, the same happens for translational models designed to deal with domain-specific data. Actually, the addition of projection matrices made KRAL+ to perform slightly worse than the pure linear translational approach used by KRAL.

In TransE and other single-hyperspace embedding approaches derived from TransE, entities from one type can induce other types of entities to form unexpected clusters when competing for space (i.e. optimizing the loss function during training). KRAL uses independent hyperspaces to embed each type of entities, in such a way that all pregnancies will form clusters that represents either the disjointness or the similarity regarding all other pregnancies, independently of other entity types, such as medications or ICD-10 codes. We believe this is a useful embedding feature when trying to represent categorized multi-relational data in specific domains.

## IV. Discussion

The findings in this study aim to identify a computational method that could support isolating the impact of each risk factor described, and improve the safety of clinical decisions before and during the pregnancy comparatively to what current protocols can actually do. We claim that KER embedding methods have the ability of learn the low-dimensional latent feature representation of the KG constituents, operate on their semantic relatedness and efficiently handle data sparsity and inconsistency issues. Subsequently, the resulting embedding representation provides the evidences to support analyzing groups of semantically related pregnancies regarding their commonly observed features.

The availability and quality of maternal and newborn health care are considered to be the “litmus test” of health systems [5]. There is an increasing interest of clinicians and federal healthcare policy makers on modeling disease risk in order to identify and analyze potential events that may negatively impact patients, lessen subsequent patient health decay, make judgments about the tolerability of the risks while considering multiple influencing factors, and reduce preventable harm and associated healthcare costs [34].

Treatment recommendations in clinical practice guidelines are supported by the evidence collected from research studies that utilize populations with highly selective sociodemographic and comorbidity-related characteristics. However, when treating complicated patients, physicians need to determine the applicability of a study to their clinical population who do not wholly align with guideline recommendations [35]. A more accurate evidence-based risk correction, supported by policies and protocols defined from data analysis, allows the health system to focus on a multi-level care stratagem, instead of only using the existing binary decision approaches (low and high risk pregnancy).

In [34], authors propose a disease risk prediction model that uses historical medical diagnoses to compound a comorbidity network to both generate a risk prediction and providing explainable rules to support clinicians to understand the resulting risk pathway. Their assumption is that diseases can progress and co-occur according to the latent relationships of underlying mechanisms and that historical medical disorders can largely affect the future onset of certain diseases. Although claiming to achieve high accurate results, they do not provide supporting evidence about the dataset in terms of balance of positive and negative cases, which makes the reported AUC scores unreliable. Moreover, they do not describe whether each diagnose was associated with a corresponding temporal reference, as the order of multiple events along the timeline can be a key factor when assessing patient risk. Finally, additional relevant socio-demographic variables were not considered, weakening the resulting model on not being able assess such inherent risks, such as those related to aging or other clinical factors (e.g. BMI).

Disease risk prediction models are preferable as longer the ability of constructing explainable rules that support understanding why and how the prediction was made. Revealing why the predicted result is made for a patient is an important condition in both heightening the patient’s trust towards the model and help physicians to choose whether or not to trust the model, combining derived information from their domain knowledge in favor of accepting or rejecting the predicted result [34]. KER can efficiently provide evidence that supports a risk assessment scale, moving from the traditional binary decision to a continuous risk scale [0,1] in order to support a multi-level risk assessment.

A study investigating 14,000 woman in China [36] points out postpartum hemorrhage, hypertension during pregnancy, diabetes, and anemia as the top observed pregnancy-related complications within that cohort, mainly correlated with delivery times, gestational weeks, and informal pregnancy examinations, whereas age at pregnancy, obesity, pre-existing medical conditions, number of pregnancies, and education were less significant, though still correlated. However, these cause patterns can vary due to demographic factors, such as when comparing high- and low-income countries [37], or when comparing maternal race and ethnicity [38].

Despite the progress in understanding the causes of early pregnancy losses, there are many mechanisms underlying maternal losses that still remain unknown, mostly related to cellular or genetic meiotic chromosomal abnormalities, that can be translated into no observed correlation with sociodemographic factors and clinical history [39]. In this sense, KER methods provide the supporting evidence to assist finding common characteristics in a K-nearest neighborhood analysis of similar cases.

Fig. 5 shows the probabilistic risk distribution of miscarriage in each maternal age range for both training and test sets used in our experiments. Although pregnancy in older women is subject to higher risk, such age-related scores are only a generalization of the cases and do not provide any concrete evidence to support patient-centric decisions.

**Fig. 5.**
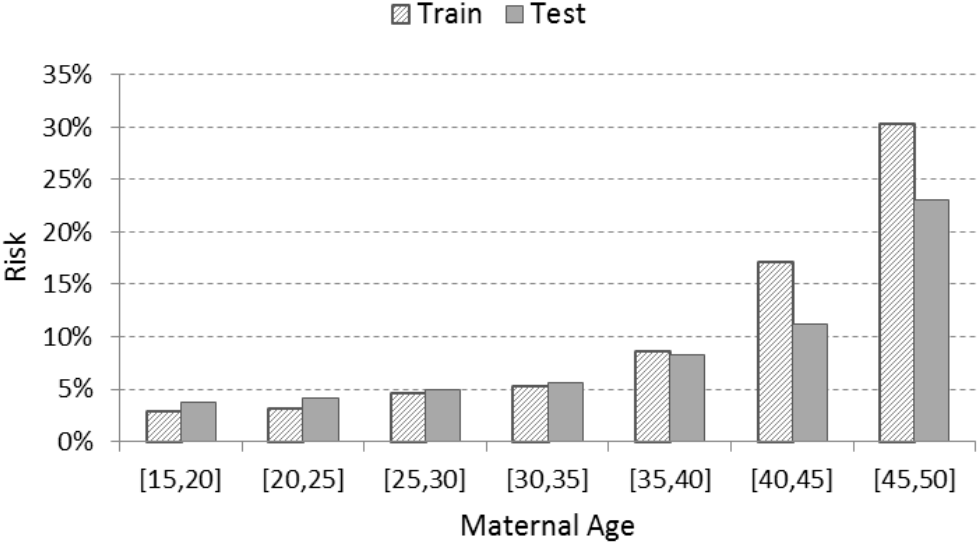
Overall probabilistic risk of miscarriage related to maternal age given by the training and test sets.

KER learns the latent feature representation of each pregnancy, making those with similar characteristics to be found in the nearby neighborhood within the hyperspace, based on their semantic relatedness. Fig. 6 shows a different scenario for the resulting risk assessment regarding each maternal age range. The average miscarriage ratios are similarly higher in all maternal ages from 35-years-old. However, pregnancy cases with the maximum ratios are observed in the range [35,40] instead of [45,50].

**Fig. 6.**
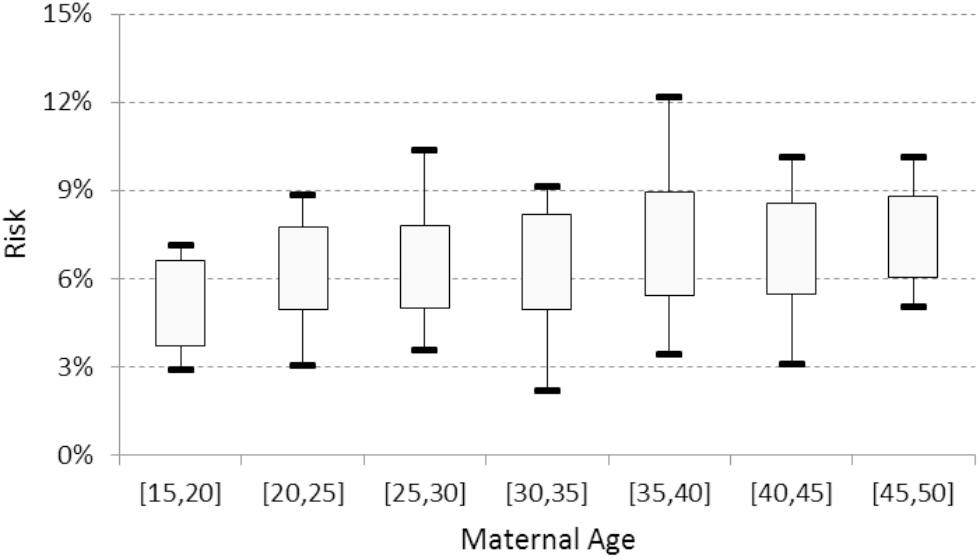
Risk of miscarriage related to maternal age resulting from embedding neighbors’ similarity – min, max, and avg*±*stdev miscarriage ratios considering up to 100 nearest training neighbors for each pregnancy in the test set.

Unlike probabilistic risk assessment approaches, KER is able to provide a distinct risk score for each pregnancy individually. Moreover, when the most similar cases are taken into account, the pairs of more relevant feature values can be analyzed in order to describe which characteristics are more relevant to the specific group of pregnancies semantically related to the assessed pregnancy. We randomly selected 15 pregnancy cases women over 30-years-old and we manually analyzed the top-100 feature values for each assessed pregnancy cluster. Table IV some of the most clinically relevant for each case. Pregnancy cases within the same maternal age can be described with very different features. For example, some pregnancy cases have higher assessed risk ratio and belong to a group of similar cases that share conditions known to be related to miscarriages (e.g. infections, smoking, and hypertension), whereas the use of Folic Acid is one of the features consistently describing those clusters corresponding to cases with lower risk ratio.

**Table IV.** Some of the most relevant features obtained from the KER approach for a sample of 15 patients.

| # | Age | Risk | Relevant features |
| --- | --- | --- | --- |
| 1 | 30-35 | 9.1% | history of psychological and psychiatric treatment preceding LMP – diagnosis of 'Adjustment disorders' (ICD-10 F43.2) 9-12 months before pregnancy – infectious disease tested in the first 8 weeks of pregnancy |
| 2 | 30-35 | 8.2% | use of Nimesulide and Ciprofloxacin in the quarter preceding pregnancy |
| 3 | 30-35 | 3.4% | Tenoxicam and Amoxicillin prescribed between 6-12 months before pregnancy – using Folic Acid during pregnancy |
| 4 | 30-35 | 2.1% | using Folic Acid |
| 5 | 35-40 | 12.1% | history of psychological and psychiatric treatment preceding LMP – use of Fluoxetine in the year before pregnancy – Amoxicillin prescribed between 6-12 months before pregnancy |
| 6 | 35-40 | 11.3% | smoker |
| 7 | 35-40 | 4.0% | dental treatment during pregnancy |
| 8 | 35-40 | 3.4% | smoker – using Folic Acid – OB-GYN visit in the first 8 weeks of pregnancy (high-risk considered) |
| 9 | 40-45 | 10.1% | Amoxicillin prescribed between 6-12 months before pregnancy – psychological treatment in the quarter preceding LMP |
| 10 | 40-45 | 9.8% | smoker – consistently using Amoxicillin in the 12 months preceding pregnancy – dental treatment during pregnancy – OB-GYN visit in the first 8 weeks of pregnancy (high-risk considered) |
| 11 | 40-45 | 3.9% | smoker – using Folic Acid |
| 12 | 40-45 | 3.0% | Amoxicillin prescribed between 6-12 months before pregnancy – using Folic Acid |
| 13 | 45-50 | 10.1% | hypertension |
| 14 | 45-50 | 8.1% | smoker – Amoxicillin prescribed between 6-12 months before pregnancy |
| 15 | 45-50 | 6.1% | smoker – history of urinary infection – diagnosis of 'Infectious Gastroenteritis' (ICD-10 A09) 3-6 months before pregnancy – psychiatric treatment in the semester preceding LMP – using Folic Acid during pregnancy |

Prenatal care before and during pregnancy significantly decreases the risk of complications that can lead both mother and developing fetus to death. Clinical decision support systems play an important role in all levels of the health system on assisting health providers in decision making that can improve the quality of prenatal care, identifying potential conditions that lead to high risk pregnancy, and contributing to improve quality of health care [40]. We noted that some of the characteristics observed in the analyzed cases are not commonly highlighted with great weight in the literature, neither addressed as main elements in guidelines. The possibility of assigning more individualized values to these factors (unlike the typical ‘high’ or ‘low’ risk), in addition to the patientcentered analysis, provides a more granular stratification of risks regarding miscarriages, which makes the interactions between physician and patient more straight and accurate.

## V. Conclusions

Health problems during pregnancy can affect mother’s health even when they were healthy before getting pregnant, and such complications may make the it a high-risk pregnancy, and can be caused by factor ranging from population demographics to clinical history.

In this study, we show how clinical knowledge embedding approaches can support risk assessment of miscarriage during pregnancy. We tested the ability of embedding approaches on both capturing the semantic correlations from multi-relational categorized pregnancy data, and we demonstrate that even simple embedding approaches that utilize domain-specific metadata are able to improve the risk evaluation both before and during the earlier pregnancy stages, comparatively to probabilistic approaches. We also show that embedding approaches are able to provide evidence about how the feature values of a test instance are related to the feature values, supporting explainability for its model prediction, in such a way that humans understand.

The way we plan to extend this work includes: (a) expanding current experiments to a wider pregnancy dataset (up to 20 years), reassessing risk year after year in order to draw the high-level picture on how clinical risk changes and is affected based on time; (b) evaluating multiple choices in terms of ontological constraints that can potentially improve the quality embedding representation of clinical data and subsequently improve risk assessment and further machine learning applications; and (c) validating how different training approaches could be effectively combined in order to simultaneously improve risk assessment of multiple clinical conditions based on an unified knowledge embedding representation approach.

## Data Availability

Dataset is not fully de-identified and it is not publicly available.

## Footnotes

1 Data controllers of the *InfoSaude* system have granted us permission to use and perform analysis on a de-identified version of this dataset, and no time limit has been set for data usage.

2 https://github.com/hextrato/KRAL

